# Determinants of Population Coverage Measurement for Key Health Indicators in Haïti

**DOI:** 10.1101/2025.03.26.25324728

**Authors:** Mickelder Kercy

**Affiliations:** Johns Hopkins University

**Keywords:** Health System, Primary Health Care, Health Management Information System, Financing, Policy, Haïti

## Abstract

**Introduction:** Health system strengthening is a prerequisite to achieving continuous and impactful improvements in population health outcomes and the sustainable development goals 3 (SDGs 3) in low and middle-income countries. This study aimed to characterize and estimate the association of health facilities’ determinants with their performance levels in measuring population coverage for key health indicators in Haïti.

**Methods:** The 2017-2018 Haïti DHS service provision assessment (SPA) data were used to conduct this study. A sample of 993 primary health care facilities was dichotomized into the institutions that measured population coverage for key health indicators and the institutions that did not measure population coverage for key health indicators. These institutions were profiled according to specific characteristics, health management information system infrastructure, and funding sources. A multivariable logistic regression model provided estimates of the association between these determinants and institutional performance level in measuring population coverage for key health indicators.

**Results:** Most primary health care facilities were health centers without bed (36.4%) and community health centers (35.4%), under the managing authority of the government (33.4%) or the private for-profit sector (30.3%), located in rural areas (63.3%), operating a health services data collection system (93.7%), reporting data at least once per month (85.5%), staffed with health services data management personnel (in-house designated 18.9%, in-house non-designated 22.8%, and outsourced 23.8%), and funded by out-of-pocket payment (79.8%) and the Ministry of Health (53.4%). However, health services data technology was only available and functioning in less than half of all health facilities. Further analysis revealed that Institutions that measured population health coverage were predominantly community health centers (43.6%, UOR = 3.21, 95% CI [2.09, 4.96], p < 0.001) and health centers without bed (32.0%, UOR = 1.41, 95% CI [0.92, 2.15], p = 0.11), managed by the government (46.7%) and mixed entities (22.4%, UOR = 0.54, 95% CI [0.36, 0.81], p = 0.003), had a data collection system in place (98.2%, UOR = 7.68, 95% CI [3.86, 15.31], p < 0.001), reported data frequently at least once per month (98.0%, UOR = 35.57%, 95% CI [15.45, 81.89], p < 0.001) and had complete data on non-communicable diseases (99.3%, UOR = 6.00, 95% CI [2.01, 17.89], p = 0.001) and antenatal care services (97.1%, UOR = 6.24, 95% CI [3.56, 10.95], p < 0.001). Data management personnel was either formal, informal or contracted in more than 70% of these institutions located primarily in rural areas. In contrast, merely.one-third of these institutions were equipped with health services data technology (UOR = 0.55, 95% CI [0.43, 0.72], p < 0.001). Top funding sources were mostly out-of-pocket or individual direct payments (81.6%), UOR = 1.29, 95% CI [0.95, 1.76], p = 0.106) and from the Ministry of Health (68.7%, UOR = 4.37, 95% CI [3.34, 5.71], p < 0.001). Comparatively and within group, differences were noted among institutions that did not measure population coverage for key health indicators in Haïti.

**Conclusion:** Measuring population coverage for key health indicators is vital for strengthening the health system in Haïti. Continued reinforcement of the national public health policy, incremental and long-term increase in government funding in health care, sustained health management information system capacity-building; and political will, commitment, and stability are warranted.

## Introduction

Achieving continuous and impactful improvements in the sustainable developmental goal 3 (SDG 3) requires the implementation, monitoring, evaluation, and commensurate investments in population health systems^1,2,3^. Population health indicators are international and country specific metrics that guide and inform the status of health system efficiency level, national health policy outcomes and the SDGs^1,2,3^. Measuring population coverage of key health indicators enables the adoption of a multipronged approach in the strategic prioritization of health financing, resources allocation, and relevant policies to ensure equitable and impactful provision and management of preventative, curable and chronic health services at the district and national levels^1,2,3^.

The health system in Haïti is a three-tiered network of university hospitals and other specialized institutions at the third level, departmental hospitals at the second level, and primary health care facilities at the first or frontline level^4^ designed to provide health care coverage to its population of 11,584,994 people^5^. Primary healthcare facilities include community hospitals serving a population size of approximately 150,000 to 250,000 people, and health centers with beds, health centers without bed and community health centers with catchment areas estimated at 10,000 people. In 2021, health expenditure per capita amounted to $57.88^6^ and accounted for 3.48% of the gross domestic product (GDP)^5^.

Notable improvements have been reported on maternal and child health indicators with the decrease in infant mortality from 74 per 1,000 live births in the 1994-1995 period to 59 per 1,000 live births in the 2016-2017 period^7^. Similarly, maternal health care improved as reflected by the substantial provision and utilization of antenatal care by 91% of women in the period 2016-2017 compared to 68% in the period 1994-1995^7^. However, the median antenatal care with four or more visits (ANC4+) coverage is lowest (64%) and the median neonatal mortality rate (NMR) is highest (32 per 1000 live births) among Latin American and Caribbean countries^8^. Over the period 2003 to 2017, cardiovascular diseases became the leading cause of burden of disease with 3,364 years of life lost (YLLs) per 100,000 people nationally in the Caribbean region^9^. Cardiovascular diseases are the top contributors of disability adjusted life-years (DALYs) among men and women aged 50 and above^9^. Pursuant to the SDG 3 target indicator 3.2, Haiti ought to achieve a reduction of under-5 mortality to 25 per 1,000 live births and maternal mortality ratio to 70 per 100,000 live births by the year 2030^4^. Premature non-communicable diseases are also expected to be reduced by one third^4^.

The current Haitian national public health policy has been framed to build on these and other reported successes with explicit focus on expanding the health system network and ensuring equitable coverage of the Essential Package of Health Services (EPHS)^10^. Since the year 2017, the Haitian Ministry of Public Health and Population (MSPP) has collaborated with international partners to adapt and implement a single interoperable health management information system using the District Health Information Systems 2 (DHIS2) software platform (version 2.33)^11^. As reported in 2021, the national health management information system or system d’information sanitaire national unique (SISNU) has been providing access to and transfer capabilities of paper and digital data to 66% of all health facilities and 273 health facilities equipped with mHealth platforms, and tracking data on HIV, COVID-19 and tuberculosis^11^. National coverage in health facilities’ linkage to this innovative platform and broad measurement of population level key indicators are expected to improve. The aim of this research study was to profile the institutions that were routinely or predisposed to measuring population coverage for key health indicators prior to the implementation of the SISNU health management information system in Haïti. Institutional profiles might be commensurate with their population coverage measurement performance post SISNU implementation; which may warrant further evaluations and strategic interventions.

## Methods

### Survey Design and Data source

Secondary data analysis was performed using data from the 2017-2018 cross-sectional DHS Service Provision Assessment (SPA)^7^. The 2017-2018 Haïti Service Provision Assessment (HSPA) is a health system facility-based survey that is nationally representative of the 10 departments in Haïti. In total, 1,007 out of 1,033 health facilities were sampled; with a response rate of 97%. Among the 1,007 health facilities included in this sample, the majority were surveyed in the West department (n=380), l’Artibonite department (n=124), and North department (n=108). The remaining 395 facilities were surveyed throughout 7 departments at a rate of 35 to 87 facilities per department. Weighting was performed to factor in the non-response rates.

### Study Variables and Measurements

#### Dependent variable

The dependent variable for this research study is *Institutions that measure population coverage for key indicators*. On the 2017-2018 HDHS SPA questionnaire, the leading and appropriate member of the personnel at each health facility was asked the following question “Do you measure population coverage for key indicators, like vaccination?”. Respondents were given the option to either affirm “yes”, “no”, or “do not know”. The latter two responses were recoded as “no”.

#### Independent variables

The variable *facility type* had initially seven response categories: 1. university hospital, 2. department hospital, 3. community reference hospital, 4. other hospitals, 5. health center with bed, 6. health center without bed and 7. dispensary/community health center. Primary health care (PHC) centers^2^ were selected and recoded as follows: (3 = 3 “Community hospitals”) (4 = 3 “community hospitals”) (5 = 4 “health centers with beds”) (6 = 5 “health centers without beds”) (7 = 6 “community health centers”). The variables *managing authority* were respectively relabeled *health facility sector* and *health facility location*. Both *health facility sector* and *urban/rural* variables retained their original categories. *Total outpatient visits for last completed calendar month* was relabeled to compute *mean outpatient visits in the previous month. Total outpatient visits for last completed calendar month* was the only continuous variable. The variable *system in place to regularly collect health services data* was relabeled *health services data collection system. The response categories “yes” and “no” were replaced with the terms “available” and “unavailable”. Facility has a functioning computer* was redefined *as health services data technology.* The three responses “no computer, yes functioning, yes but not functioning” for this variable were redefined as “unavailable, functioning, not functioning). The variable *frequency of reports containing health services data* was relabeled *health services data reporting frequency.* This variable had six categories (0-5), of which three (1, 4 and 5) were retranslated from the French questionnaire to English, as follows: 0. never, 1. at least once per month, 2. every 2-3 months, 3. every 4-6 months, 4. less than once every six months, and 5. no answer. These categories were then recoded (0 5 = 0 “never”) (1 = 1 “At least once per month”) (2 3 4 = 2 “Once every 2-6 months”). The variable *designated person responsible for health services data* was replaced with the term *health services data management personnel*, and all categories (0. no, no designated person, 1. yes, data manager/HMIS person/statis, 2. yes, facility in-charge, and 3. yes, other service provider) were renamed respectively (0. none, 1. in house – specified, 2. in house – non specified, and 3. Outsourced). The response variables “no” and “yes” for data on non-communicable diseases and antenatal care services were relabeled “unavailable” and “available”, respectively. Twenty-four responses “don’t know” for *total outpatient visits for last completed calendar month* were substituted with the numerical value zero to match the survey responses from 28 other health facilities. All other variables retained their core definitions and response categories “no” and “yes”.

#### Ethical consideration

Permission was requested and obtained in May 2024 from the Demographic Health Survey Office to utilize the 2017-2018 HDHS SPA dataset. As stipulated on the DHS SPA questionnaire, a leading personnel member per health facility provided oral informed consent prior to conducting the service provision assessment portion.

#### Data analysis

The total number of observations (n=993) was estimated for the dependent and independent variables in this study.

Descriptive statistics of the independent variables were computed according to the binary dependent variable. The Rao and Scott second-order corrected Chi-square was used to analyze the relationships between these variables. Outpatient volume was tabulated according to the health facility characteristics.

Statistical models were developed to assess and evaluate any existing associations between the reported and documented characteristics of health facilities, health management information systems, presence of data on non-communicable diseases and antenatal care, and funding sources with the performance level of institutions in measuring population coverage for key indicators in Haïti. The final multivariable statistical analysis was performed including all variables according to the assumption of a survey design with weight using the svy package.

All statistical analyses were performed using the software package Stata version 18, StataCorp^12^.

## Results

### Sample Description

Overall, measuring population coverage for key health indicators was performed in 56.29% (n = 559) of the 993 primary health care facilities. Health centers without bed (36.4%) and community health centers (35.4%) comprised the majority of the weighted sample of primary health care facilities (n = 993). Approximately, 33.4% of primary health care facilities were managed by the government and 30.3% were privately-owned for-profit institutions (Table1). Nearly two-thirds of health facilities were in rural areas.

Health services data collection systems were in operation in 93.7% of the total sampled primary health care institutions (Table 1). In this same sample, health services data technology (computer) was either not available (54.4%), functioning (40.0%), or not functioning (5.6%). Estimated health services data reporting frequencies were as follows: never (12.4%), at least once per month (85.5%) and once every 2-6 months (2.1%). Across all primary health care facilities, health services data management personnel did not exist (34.5%), had been specified (18.9%), had not been specified (22.8%) and had been outsourced (23.8%).

**Table 1.**
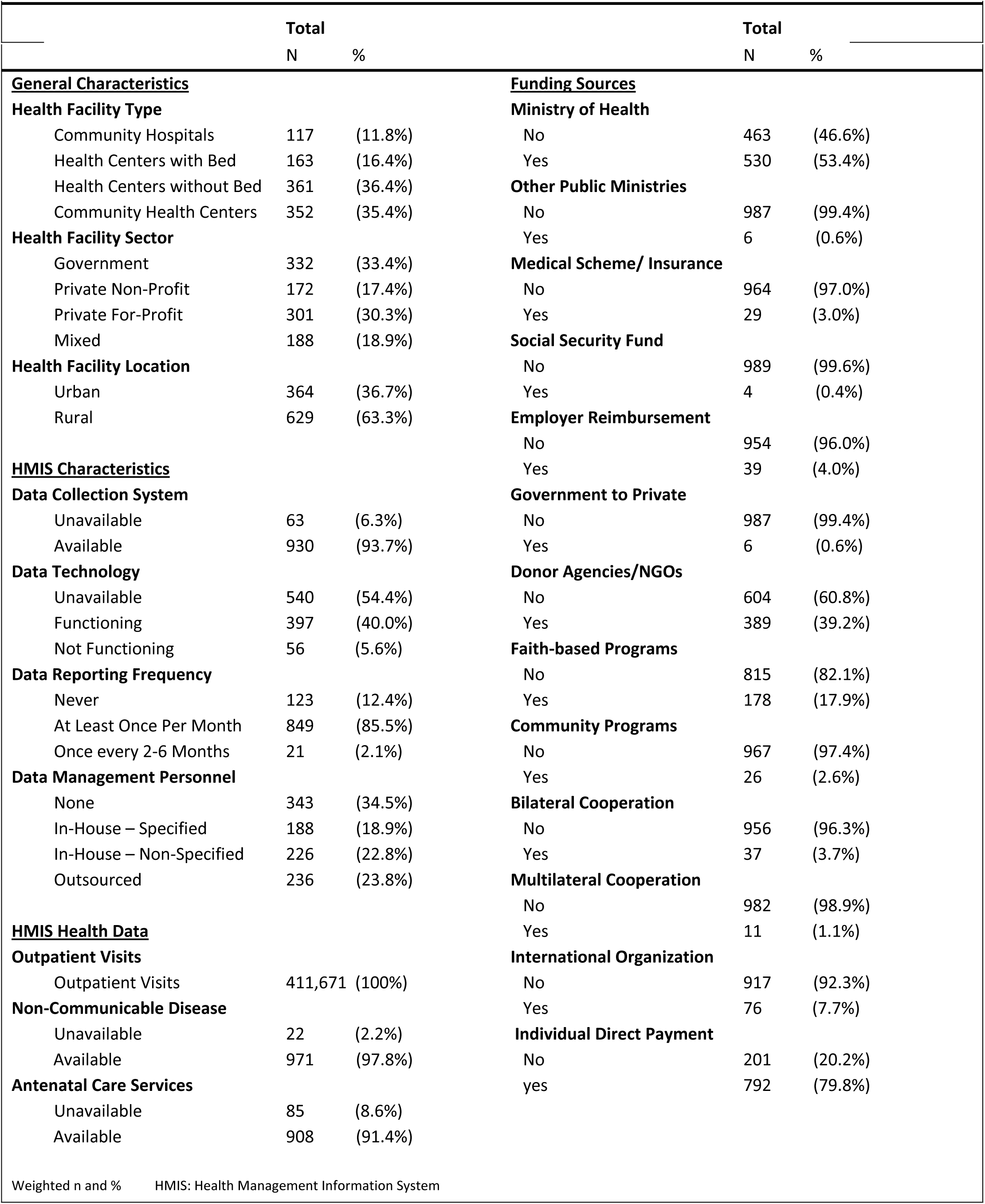
General Distribution of Primary Health Care Facility by Characteristics and Funding Sources in Haïti

|  | N | % |  | N | % |
| --- | --- | --- | --- | --- | --- |
| **<u>General Characteristics</u>** |  |  | **<u>Funding Sources</u>** |  |  |
| **Health Facility Type** |  |  | **Ministry of Health** |  |  |
| Community Hospitals | 117 | (11.8%) | No | 463 | (46.6%) |
| Health Centers with Bed | 163 | (16.4%) | Yes | 530 | (53.4%) |
| Health Centers without Bed | 361 | (36.4%) | **Other Public Ministries** |  |  |
| Community Health Centers | 352 | (35.4%) | No | 987 | (99.4%) |
| **Health Facility Sector** |  |  | Yes | 6 | (0.6%) |
| Government | 332 | (33.4%) | **Medical Scheme/ Insurance** |  |  |
| Private Non-Profit | 172 | (17.4%) | No | 964 | (97.0%) |
| Private For-Profit | 301 | (30.3%) | Yes | 29 | (3.0%) |
| Mixed | 188 | (18.9%) | **Social Security Fund** |  |  |
| **Health Facility Location** |  |  | No | 989 | (99.6%) |
| Urban | 364 | (36.7%) | Yes | 4 | (0.4%) |
| Rural | 629 | (63.3%) | **Employer Reimbursement** |  |  |
| **<u>HMIS Characteristics</u>** |  |  | No | 954 | (96.0%) |
| **Data Collection System** |  |  | Yes | 39 | (4.0%) |
| Unavailable | 63 | (6.3%) | **Government to Private** |  |  |
| Available | 930 | (93.7%) | No | 987 | (99.4%) |
| **Data Technology** |  |  | Yes | 6 | (0.6%) |
| Unavailable | 540 | (54.4%) | **Donor Agencies/NGOs** |  |  |
| Functioning | 397 | (40.0%) | No | 604 | (60.8%) |
| Not Functioning | 56 | (5.6%) | Yes | 389 | (39.2%) |
| **Data Reporting Frequency** |  |  | **Faith-based Programs** |  |  |
| Never | 123 | (12.4%) | No | 815 | (82.1%) |
| At Least Once Per Month | 849 | (85.5%) | Yes | 178 | (17.9%) |
| Once every 2-6 Months | 21 | (2.1%) | **Community Programs** |  |  |
| **Data Management Personnel** |  |  | No | 967 | (97.4%) |
| None | 343 | (34.5%) | Yes | 26 | (2.6%) |
| In-House – Specified | 188 | (18.9%) | **Bilateral Cooperation** |  |  |
| In-House – Non-Specified | 226 | (22.8%) | No | 956 | (96.3%) |
| Outsourced | 236 | (23.8%) | Yes | 37 | (3.7%) |
| **<u>HMIS Health Data</u>** |  |  | **Multilateral Cooperation** |  |  |
| **Outpatient Visits** |  |  | No | 982 | (98.9%) |
| Outpatient Visits | 411,671 | (100%) | Yes | 11 | (1.1%) |
| **Non-Communicable Disease** |  |  | **International Organization** |  |  |
| Unavailable | 22 | (2.2%) | No | 917 | (92.3%) |
| Available | 971 | (97.8%) | Yes | 76 | (7.7%) |
| **Antenatal Care Services** |  |  | **Individual Direct Payment** |  |  |
| Unavailable | 85 | (8.6%) | No | 201 | (20.2%) |
| Available | 908 | (91.4%) | yes | 792 | (79.8%) |

Weighted n and % HMIS: Health Management Information System

Health care financing from the Ministry of Health covered 53.4% of all primary health care facilities (Table 1). Donor agencies/NGOs contributed financially to 39.1% of all these institutions. Faith-based programs provided funding to 17.9% of all health facilities. 79.8% of all health facilities received individual direct payment for health care services. Additional funding sources included other government ministries, medical schemes / insurance, social security funds, employer reimbursement, government to private, bilateral cooperation, multilateral cooperation, and international organization, with levels varying from 0.4% to 7.7%.

Mean outpatient visits for the month preceding the health facility service provision assessment have been estimated (see Table A in Appendix). The mean number of outpatient visits was 1,045, 95% CI [795, 1296] for community hospitals and lowest at 175, 95% CI [143, 208]) for health centers without bed. Mean outpatient visits varied between 376 (95% CI [293, 460] and 470 (95% CI [355, 585], respectively at health facilities under governmental and mixed entities. There were 562 outpatient visits (95% CI [457, 668] reported in urban areas and 329 outpatient visits (95% CI [277, 381]) in rural areas. Primary health care institutions that measured population coverage for key indicators had on average 312 outpatient visits (95% CI [254, 369]) compared to those that did not (*x̄* = 2283, 95% CI [415, 573]). Further differences were noted and reported on Table B in the Appendix section.

When comparing the institutions that measured population coverage for key health indicators with those that do not, community health centers (43.6% vs 25.2%) and health centers without bed (32.0% vs 41.9%) ranked highest but reversely (Table 2). In unadjusted analyses, institutions that measured population coverage for key health indicators were three times more likely to be community health centers OR = 3.21, 95% CI [2.09, 4.96], p < 0.001). Less than half (46.7%) of institutions that measured population coverage for health indicators were managed by the government. Comparatively, 45.4% and 23.7% of the institutions that did not measure population coverage for key health indicators were respectively privately-owned for-profit and privately-owned non-profit health facilities. A high proportion of institutions that measured population coverage for key health indicators were found in rural areas (71.4%, OR = 2.21, 95% CI [1.70, 2.87], p < 0.001). The percentages of institutions that did not measure population coverage for key health indicators were 47.0% for urban and 53.0% for rural areas. The institutions that did not measure population coverage for key health indicators showed the highest mean number of outpatient visits for community hospitals (*x̄* = 602, 95% CI [386, 817]) and the least mean number for community health centers.(Figure 1). The reverse was true for community health centers (x̄ = 117, 95% CI [86, 147]. Similar patterns were observed when multilevel mean outpatient visits per facility type in urban versus rural areas were estimated according to the performance level of institutional measurement of population coverage for key health indicators (see appendix section, Figures 3). Mean outpatient visits were highest in community hospitals that did not measure population coverage for key indicators and who were under the authority of mixed entities (*x̄* = 1957, 95% CI [322, 3591]). In contrast, mean outpatient visits were highest (*x̄* = 1671, 95% CI [-1095, 2247]) at community hospitals under governmental authority that performed measurement of population coverage for key health indicators (see appendix section, Figure 4)

**Table 2.** Association of Primary Health Care Facility Characteristics with Institutional Measurement of Population Coverage for Key Health Indicators in Haïti

|  | Institutional Measurement of Population Coverage for Key Indicators |  |  |  |  |  |
| --- | --- | --- | --- | --- | --- | --- |
|  | No |  | Yes |  | Crude Odds Ratio |  |
|  | n | % | n | % | OR | [95% CI] |
| **<u>General Characteristics</u>** |  |  |  |  |  |  |
| **Health Facility Type** |  |  |  |  |  |  |
| Community Hospitals | 69 | (15.9%) | 48 | (8.5%) | *Reference* |  |
| Health Centers with Bed | 74 | (17.1%) | 89 | (15.9%) | 1.71* | [1.06, 2.76] |
| Health Centers without Bed | 182 | (41.9%) | 179 | (32.0%) | 1.41 | [0.92, 2.15] |
| Community Health Centers | 109 | (25.1%) | 243 | (43.6%) | 3.21** | [2.09, 4.96] |
| **Health Facility Sector** |  |  |  |  |  |  |
| Government | 71 | (16.4%) | 261 | (46.7%) | *Reference* |  |
| Private Non-Profit | 103 | (23.7%) | 69 | (12.3%) | 0.18*** | [0.12, 0.27] |
| Private For Profit | 197 | (45.4%) | 104 | (18.6%) | 0.14*** | [0.10, 0.20] |
| Mixed | 63 | (14.5%) | 125 | (22.4%) | 0.54** | [0.36, 0.81] |
| **Health Facility Location** |  |  |  |  |  |  |
| Urban | 204 | (47.0%) | 160 | (28.6%) | *Reference* |  |
| Rural | 230 | (53.0%) | 399 | (71.4%) | 2.21*** | [1.70, 2.87] |
| **<u>HMIS Characteristics</u>** |  |  |  |  |  |  |
| **Data Collection System** |  |  |  |  |  |  |
| Unavailable | 53 | (12.2%) | 10 | (1.8%) | *Reference* |  |
| Available | 381 | (87.8%) | 549 | (98.2%) | 7.68*** | [3.86, 15.31] |
| **Data Technology** |  |  |  |  |  |  |
| Unavailable | 202 | (46.5%) | 338 | (60.5%) | *Reference* |  |
| Functioning | 206 | (47.5%) | 191 | (34.2%) | 0.55*** | [0.43, 0.72] |
| Not Functioning | 26 | (6.0%) | 30 | (5.3%) | 0.69 | [0.39, 1.19] |
| **Data Reporting Frequency** |  |  |  |  |  |  |
| Never | 117 | (27.0%) | 6 | (1.1%) | *Reference* |  |
| At Least Once Per Month | 301 | (69.3%) | 548 | (98.0%) | 35.57*** | [15.45, 81.89] |
| Once Every 2-6 Months | 16 | (3.7%) | 5 | (0.9%) | 6.13** | [1.67, 22.48] |
| **Data Management Personnel** |  |  |  |  |  |  |
| None | 183 | (42.2%) | 160 | (28.6%) | *Reference* |  |
| In-House Specified | 64 | (14.7%) | 124 | (22.2%) | 2.21*** | [1.53, 3.20] |
| In-House Non-Specified | 70 | (16.1%) | 156 | (27.9%) | 2.56*** | [1.80, 3.64] |
| Outsourced | 117 | (27.0%) | 119 | (21.3%) | 1.16 | [0.83, 1.62] |
| **<u>HMIS Health Data</u>** |  |  |  |  |  |  |
| **Outpatient Visits** |  |  |  |  |  |  |
| Outpatient Visits | 135,088 | (32.81%) | 276,583 | (67.19%) | 1.00** | [1.00, 1.00] |
| **Non-Communicable Disease** |  |  |  |  |  |  |
| Unavailable | 18 | (4.1%) | 4 | (0.7%) | *Reference* |  |
| Available | 416 | (95.9%) | 555 | (99.3%) | 6.00** | [2.01, 17.89] |
| **Antenatal Care Services** |  |  |  |  |  |  |
| Unavailable | 69 | (15.9%) | 16 | (2.9%) | *Reference* |  |
| Available | 365 | (84.1%) | 543 | (97.1%) | 6.24*** | [3.56, 10.95] |
| Weighted n and % Weighted Crude Odds Ratio and 95% CI * p < 0.05 ** p < 0.01 *** p < 0.001 |  |  |  |  |  |  |

**Figure 1.**
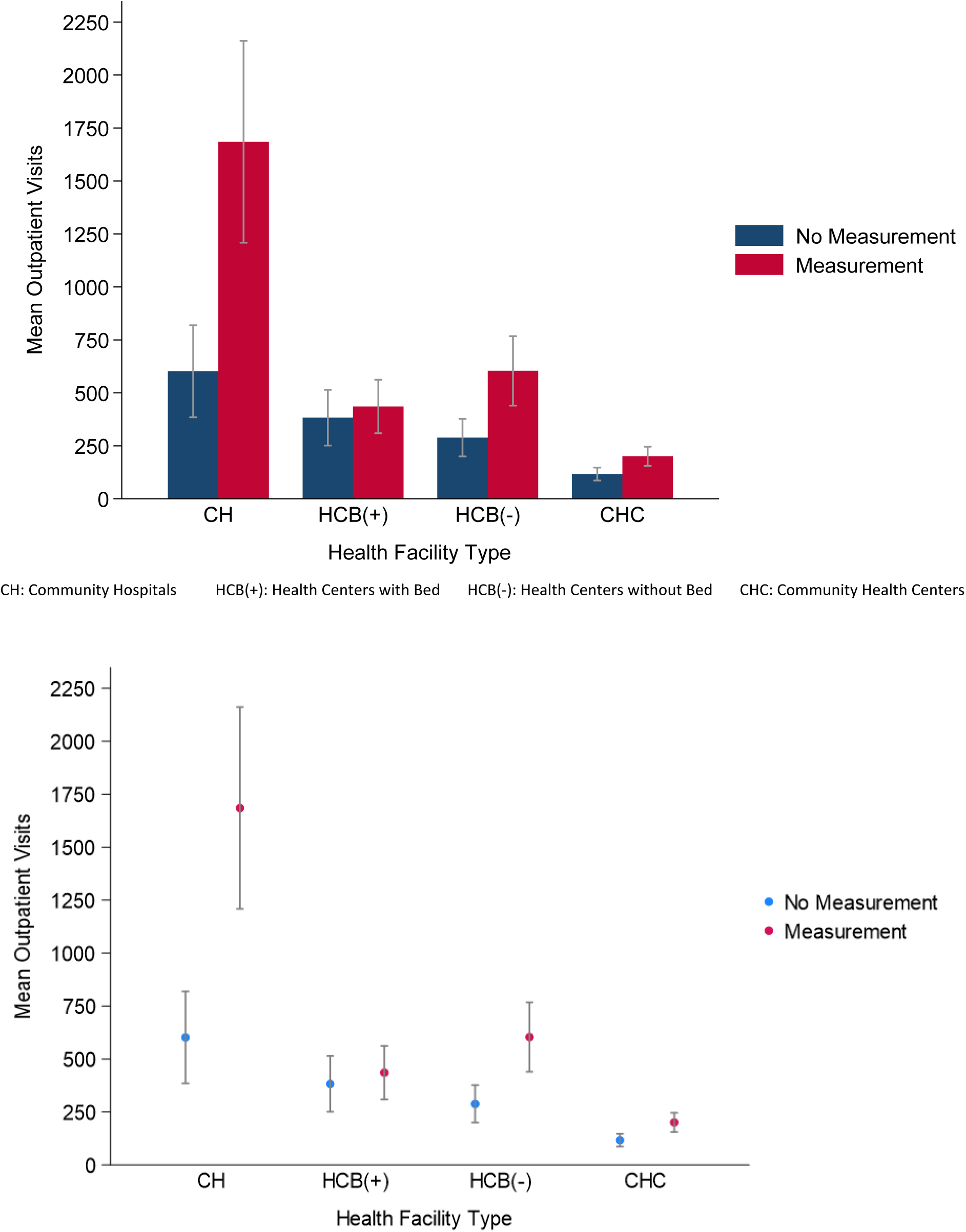
Mean Outpatient Visits in the Previous Month Per Primary Health Care Facility Type according to Institutional Measurement Status of Population Coverage for Key Health Indicators in Haïti

**Figure 2.**
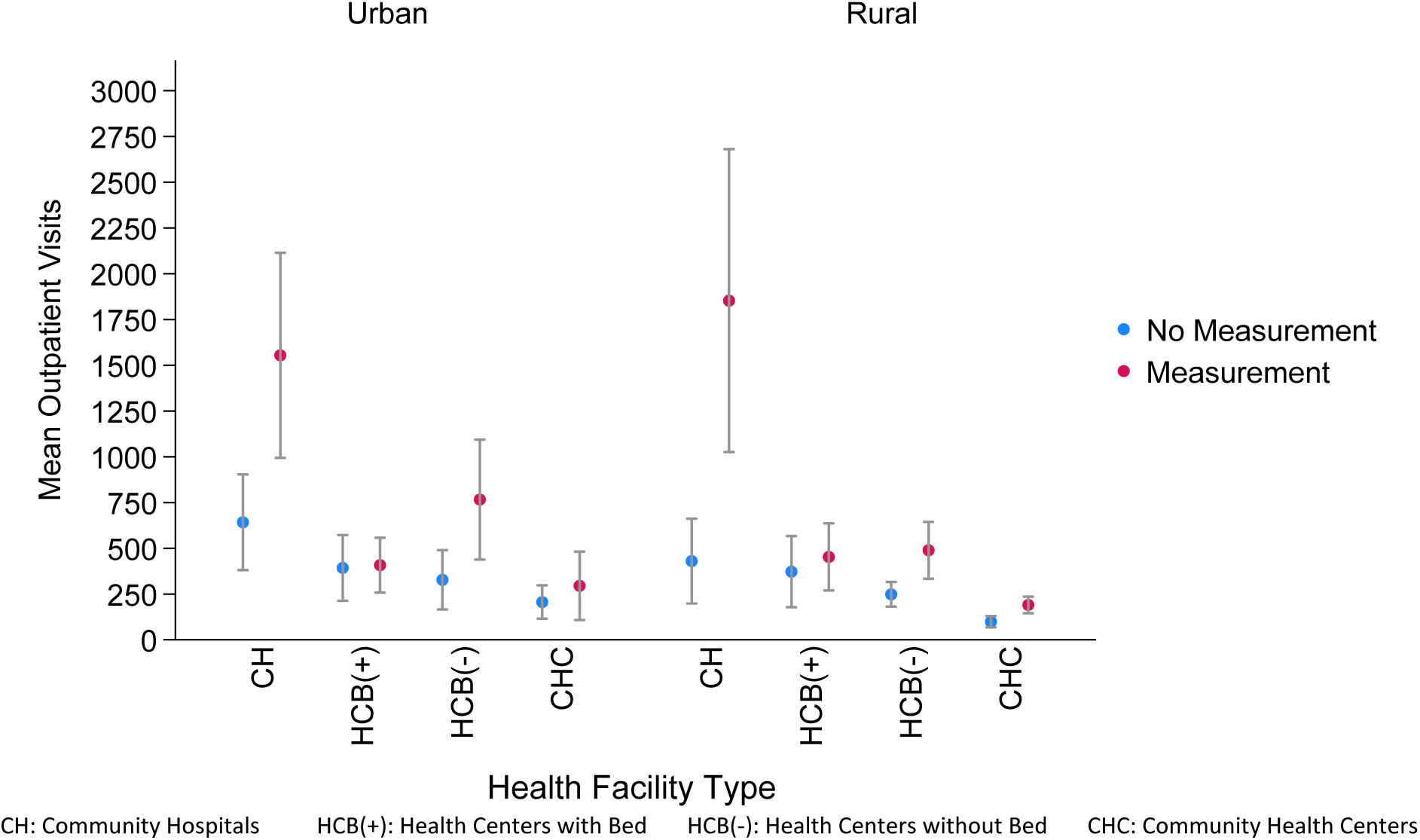
Mean Outpatient Visits In the Previous Month Per Primary Health Care Facility Type in Urban and Rural Areas according to Institutional Measurement Status of Population Coverage for Key Health Indicators in Haïti

**Figure 3.**
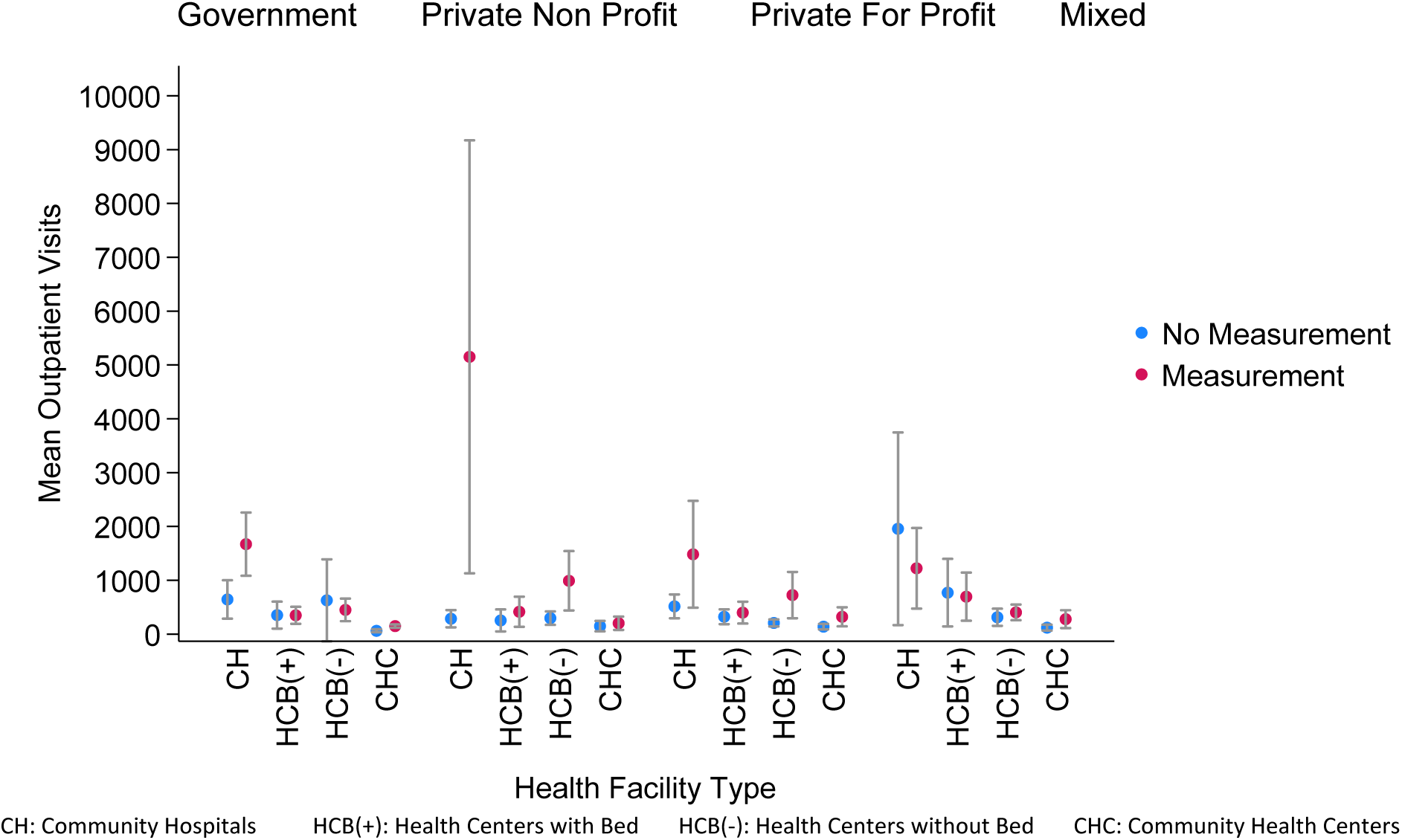
Mean Outpatient Visits In the Previous Month Per Primary Health Care Facility Type by Management Authority Type according to Institutional Measurement Status of Population Coverage for Key Health Indicators in Haïti

Health service data collection system was available in 98.2% of institutions measuring population coverage for key health indictors with an odds ratio of 7.68 (95% CI [3.86, 15.31], p < 0.001). Health services data technology (computer) was unavailable in many cases (60.5%) and when available the estimated odds of functionality was 55% (95% CI [0.43, 0.72], p < 0.001) in 34.2% of institutions measuring population coverage for key health indicators (Table 2). The frequency of health services data reporting was at least once or more per month among 98% of institutions that measured population coverage of key health indicators, OR = 35.57%, 95% CI [15.45, 81.89], p < 0.001. This frequency was similar among 69.3% of institutions that did not measure population coverage for key health indicators. Approximately a quarter of institutions that measured population coverage for key health indicators had no health services data management personnel, had designated a specific member (OR = 2.21, 95% CI [1.53, 3.20], p < 0.001), had a nonspecific member (OR = 2.56, 95% CI [1.80, 3.64], p < 0.001), or outsourced their data management service (OR = 1.16, 95% CI [0.83, 1.62], p = 0.374). Nearly a double percentage of absence of data management personnel was observed among institutions that did not measure population coverage for key health indicators. Complete health services data were reported available (97.8%) for non-communicable diseases and available (91.4%) for antenatal care services. Among institutions that measure population coverage for key health indicators, the trend was similar for health services data on non-communicable diseases (OR = 6.00, 95% CI [2.01, 17.89], p = 0.001) and antenatal care services (OR = 6.24, 95% CI [3.56, 10.95], p < 0.001).

Health care financing sources ranked similarly when comparing institutions that measured population coverage for key health indicators with those that did not (Table 3 and Table C in Appendix). The top five sources of funding in the former category were individual direct payments (81.6%, OR = 1.29, 95% CI [0.95, 1.76], p = 0.106), Ministry of Health (68.7%, OR = 4.37, 95% CI [3.34, 5.71], p < 0.001), donor agencies / NGOs (46.2%, OR = 1.98, 95% CI [1.52, 2.58], p < 0.001), faith-based programs (14.0%, OR = 0.54, 95% CI [0.39, 0.76], p < 0.001), and international organizations (9.7%, OR = 2.00, 95% CI [1.20, 3.35], p = 0.106).

**Table 3.** Top 5 Funding Sources Association with Institutional Measurement of Population Coverage for Key Health Indicators in Haïti

| **Table 3. Top 5 Funding Sources Association with Institutional Measurement of Population Coverage for Key Health Indicators in Haiti** |  |  |  |  |  |  |
| --- | --- | --- | --- | --- | --- | --- |
|  | **Institutional Measurement of Population Coverage for Key Indicators** |  |  |  |  |  |
|  | **No** |  | **Yes** |  | **Crude Odds Ratio** |  |
|  | n | % | n | % | OR | [95% CI] |
| **<u>Funding Sources</u>** |  |  |  |  |  |  |
| **Ministry of Health** |  |  |  |  |  |  |
| No | 288 | (66.4%) | 175 | (31.3%) | *Reference* |  |
| Yes | 146 | (33.6%) | 384 | (68.7%) | 4.37*** | [3.34, 5.71] |
| **Donor Agencies / NGOs** |  |  |  |  |  |  |
| No | 303 | (69.8%) | 301 | (53.8%) | *Reference* |  |
| Yes | 131 | (30.2%) | 258 | (46.2%) | 1.98*** | [1.52, 2.58] |
| **Faith-based Programs** |  |  |  |  |  |  |
| No | 334 | (77.0%) | 481 | (86.0%) | *Reference* |  |
| Yes | 100 | (23.0%) | 78 | (14.0%) | 0.54*** | [0.39, 0.76] |
| **International Organization** |  |  |  |  |  |  |
| No | 412 | (94.9%) | 505 | (90.3%) | *Reference* |  |
| Yes | 22 | (5.1%) | 54 | (9.7%) | 2.00** | [1.20, 3.35] |
| **Individual Direct Payment** |  |  |  |  |  |  |
| No | 98 | (22.6%) | 103 | (18.4%) | *Reference* |  |
| Yes | 336 | (77.4%) | 456 | (81.6%) | 1.29 | [0.95, 1.76] |
| Weighted n and % Weighted Crude Odds Ratio and 95% CI * p < 0.05 ** p < 0.01 *** p < 0.001 HMIS: Health Management Information System |  |  |  |  |  |  |

The Rao and Scott second-order corrected Chi-square was statistically significantly different (p < 0.001) between the two groups of institutions for all independent variables, with some exceptions for funding sources (other public ministries, social security fund, government to private, community programs, multilateral cooperation, and individual direct payment) (Table 5).

### Multivariable Logistic Regression Model

Table 4 shows estimates of the likelihood of primary health care institutions measuring versus not measuring population coverage for key health indicators controlling for all other variables. Based on the health facility types, compared to community hospitals, community health centers had 3.50 times higher odds to be primary health care institutions that measured population coverage for key health indicators (95% CI [1.75, 7.01], *p* < 0.001). This association was also statistically significant for health centers with or without bed.

When accounting for the managing authorities and considering the government sector as a reference group, primary health care facilities in the private sector with the non-profit status were 47% more likely to measure population coverage for key health indicators (95% CI [0.29, 0.77], *p* < 0.01). Health facilities managed by private entities for profit were 37% more likely to measure population coverage for key health indicators (95% CI [0.24, 0.58], *p* < 0.001). The odds ratio of measuring population coverage for key health indicators were 0.90 but not statistically significant for health facilities with mixed managing authorities (*p* = 0.68).

The odds of measuring population coverage for key health indicators were not statistically different when comparing primary health care facilities located in rural (OR = 1.07, , 95% CI [0.74, 1.54]) with those located in urban areas.

In regard to the health management information system, the odds of institutions measuring population coverage for key health indicators were not statistically significantly different when a data collection system was available (OR = 1.57 (95% CI [0.46, 5.34]). Compared to institutions with no data technology (computer), the likelihood of institutions measuring population coverage for key health indicators was 58% higher (95% CI [0.37, 0.90], *p* < 0.05) when the technology was available and functioning. The odds of institutions measuring population coverage for key health indicators were not statistically significant when the available technology was not functional.

Primary health care Institutions that measured population coverage for key health indicators were more likely to report data reporting frequency of at least once per month (OR = 17.43,95% CI [6.10, 49.81], *p* < 0.001) than never. The odds of institutions measuring population coverage for key health indicators with data reporting frequency of once every 2-6 months was 4.41 (95% CI [1.02, 19.06], *p* = 0.05). Compared to institutions with no data management personnel, the odds of institutions measuring population coverage for key health indicators was 1.84 (95% CI [1.05, 3.24], *p* < 0.01) for health facilities with designated personnel, 1.53 (95% CI [0.98, 2.37], *p* = 0.06) for health facilities with non-designated personnel, and 1.07 (95% CI [0.68, 1.70], *p* = 0.77) for health facilities that outsourced their data management personnel.

Estimates were provided for selective health management information system data, notably outpatient visits, non-communicable diseases, and antenatal care services. The odds of institutions measuring or not measuring population coverage for key health indicators were statistically significantly different (*p* = 0.004) for every one unit increase in outpatient visits. The estimated odds that primary health care institutions measure population coverage for key health indicators weres 3.41, 95% CI [0.76, 15.23], *p* = 0.11 when data were available on non-communicable diseases and 3.42, 95% CI [1.73, 6.74], *p* < 0.001) for antenatal care services.

The odds of institutions measuring population coverage for key health indicators was statistically significantly higher when funding sources were the ministry of health (OR = 1.99, 95% CI [1.41, 2.81], *p* < 0.001), social security fund (OR = 24.19, 95% CI [5.50, 115.28], *p* < 0.001), government to private (OR = 76.20, 95% CI [7.42, 782.90], p < 0.001), and donor agencies/NGOs (OR = 1.45, 95% CI [1.02, 2.06], *p* = 0.04), Faith-based programs were significantly associated with lower odds of population-level measurement (OR = 0.48, 95% CI [0.31, 0.73], *p* < 0.01).

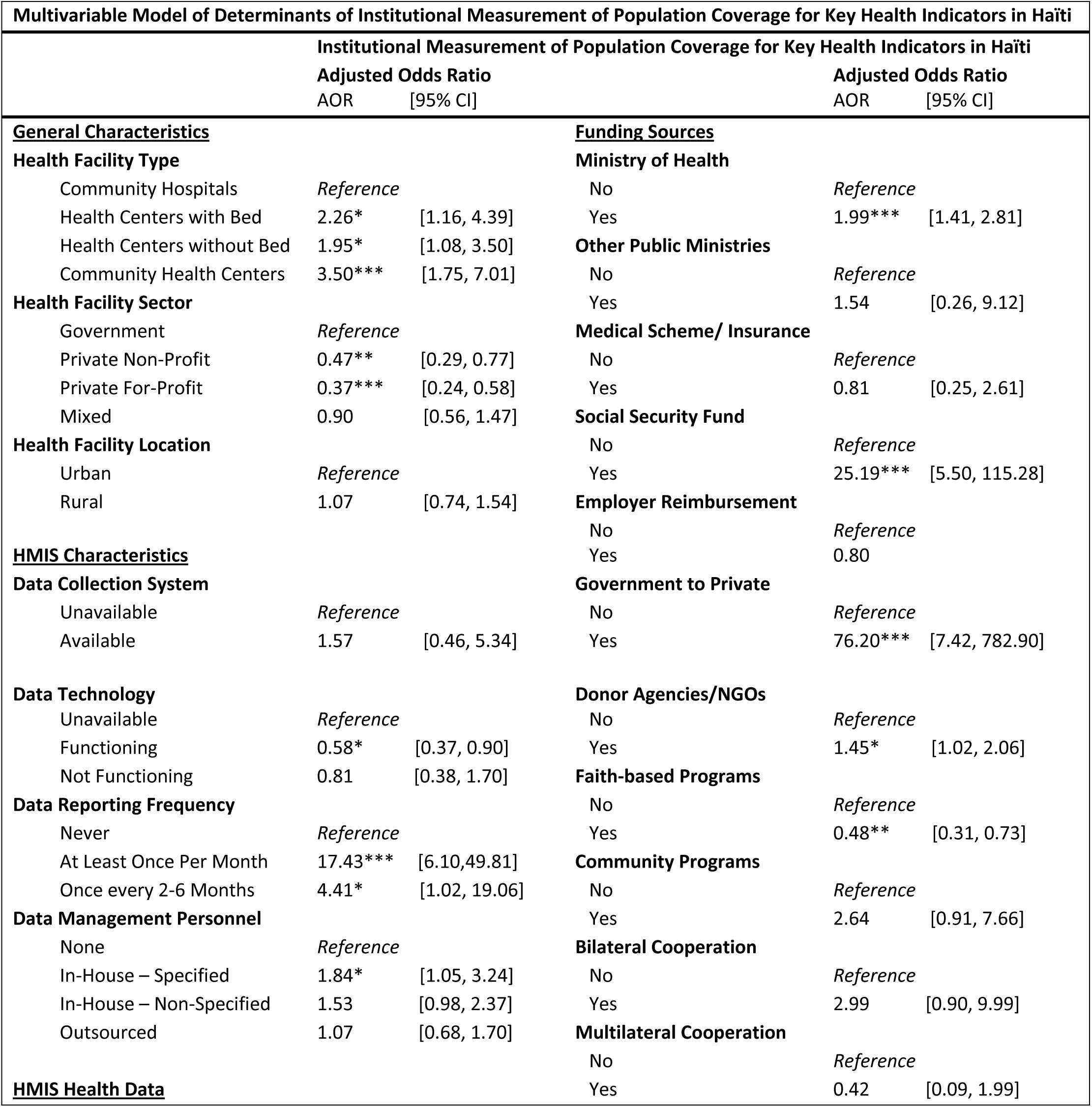
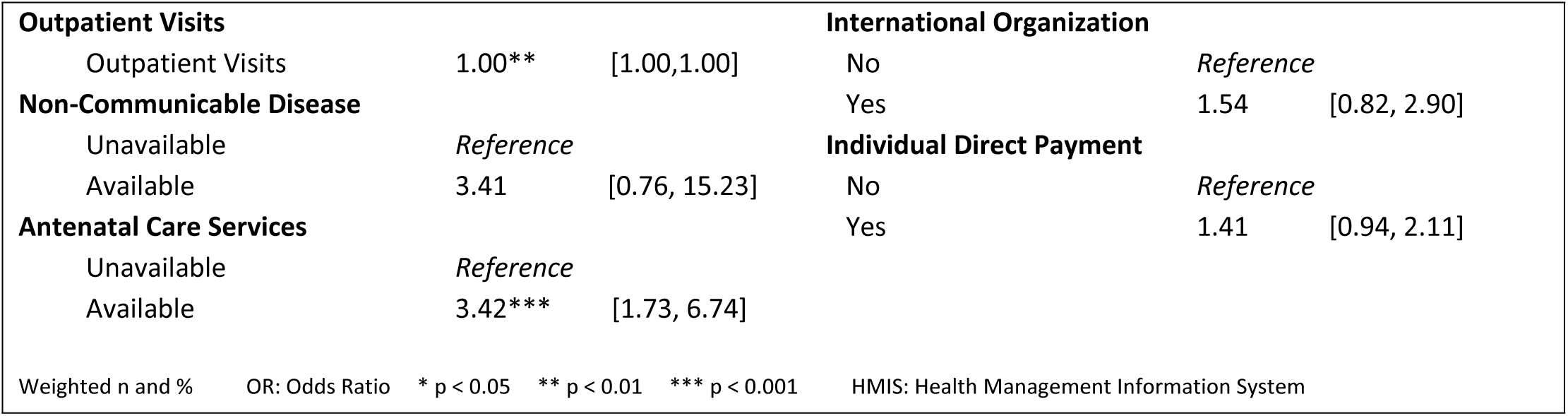

| Multivariable Model of Determinants of Institutional Measurement of Population Coverage for Key Health Indicators in Haïti |  |  |  |  |
| --- | --- | --- | --- | --- |
| Institutional Measurement of Population Coverage for Key Health Indicators in Haïti |  |  | Adjusted Odds Ratio |  |
|  |  |  | AOR | [95% CI] |
| **General Characteristics** |  |  |  |  |
| **Health Facility Type** |  |  |  |  |
| Community Hospitals |  |  | Reference |  |
| Health Centers with Bed |  |  | 2.26* | [1.16, 4.39] |
| Health Centers without Bed |  |  | 1.95* | [1.08, 3.50] |
| Community Health Centers |  |  | 3.50*** | [1.75, 7.01] |
| **Health Facility Sector** |  |  |  |  |
| Government |  |  | Reference |  |
| Private Non-Profit |  |  | 0.47** | [0.29, 0.77] |
| Private For-Profit |  |  | 0.37*** | [0.24, 0.58] |
| Mixed |  |  | 0.90 | [0.56, 1.47] |
| **Health Facility Location** |  |  |  |  |
| Urban |  |  | Reference |  |
| Rural |  |  | 1.07 | [0.74, 1.54] |
| **HMIS Characteristics** |  |  |  |  |
| **Data Collection System** |  |  |  |  |
| Unavailable |  |  | Reference |  |
| Available |  |  | 1.57 | [0.46, 5.34] |
| **Data Technology** |  |  |  |  |
| Unavailable |  |  | Reference |  |
| Functioning |  |  | 0.58* | [0.37, 0.90] |
| Not Functioning |  |  | 0.81 | [0.38, 1.70] |
| **Data Reporting Frequency** |  |  |  |  |
| Never |  |  | Reference |  |
| At Least Once Per Month |  |  | 17.43*** | [6.10, 49.81] |
| Once every 2-6 Months |  |  | 4.41* | [1.02, 19.06] |
| **Data Management Personnel** |  |  |  |  |
| None |  |  | Reference |  |
| In-House – Specified |  |  | 1.84* | [1.05, 3.24] |
| In-House – Non-Specified |  |  | 1.53 | [0.98, 2.37] |
| Outsourced |  |  | 1.07 | [0.68, 1.70] |
| **HMIS Health Data** |  |  |  |  |
| **Funding Sources** |  |  |  |  |
| **Ministry of Health** |  |  |  |  |
| No |  |  | Reference |  |
| Yes |  |  | 1.99*** | [1.41, 2.81] |
| **Other Public Ministries** |  |  |  |  |
| No |  |  | Reference |  |
| Yes |  |  | 1.54 | [0.26, 9.12] |
| **Medical Scheme/ Insurance** |  |  |  |  |
| No |  |  | Reference |  |
| Yes |  |  | 0.81 | [0.25, 2.61] |
| **Social Security Fund** |  |  |  |  |
| No |  |  | Reference |  |
| Yes |  |  | 25.19*** | [5.50, 115.28] |
| **Employer Reimbursement** |  |  |  |  |
| No |  |  | Reference |  |
| Yes |  |  | 0.80 |  |
| **Government to Private** |  |  |  |  |
| No |  |  | Reference |  |
| Yes |  |  | 76.20*** | [7.42, 782.90] |
| **Donor Agencies/NGOs** |  |  |  |  |
| No |  |  | Reference |  |
| Yes |  |  | 1.45* | [1.02, 2.06] |
| **Faith-based Programs** |  |  |  |  |
| No |  |  | Reference |  |
| Yes |  |  | 0.48** | [0.31, 0.73] |
| **Community Programs** |  |  |  |  |
| No |  |  | Reference |  |
| Yes |  |  | 2.64 | [0.91, 7.66] |
| **Bilateral Cooperation** |  |  |  |  |
| No |  |  | Reference |  |
| Yes |  |  | 2.99 | [0.90, 9.99] |
| **Multilateral Cooperation** |  |  |  |  |
| No |  |  | Reference |  |
| Yes |  |  | 0.42 | [0.09, 1.99] |

|  |  |  |  |  |  |
| --- | --- | --- | --- | --- | --- |
| **Outpatient Visits** |  |  | **International Organization** |  |  |
| Outpatient Visits | 1.00** | [1.00,1.00] | No | Reference |  |
| **Non-Communicable Disease** |  |  | Yes | 1.54 | [0.82, 2.90] |
| Unavailable | Reference |  | **Individual Direct Payment** |  |  |
| Available | 3.41 | [0.76, 15.23] | No | Reference |  |
| **Antenatal Care Services** |  |  | Yes | 1.41 | [0.94, 2.11] |
| Unavailable | Reference |  |  |  |  |
| Available | 3.42*** | [1.73, 6.74] |  |  |  |
| Weighted n and % | OR: Odds Ratio | * p < 0.05 ** p < 0.01 *** p < 0.001 | HMIS: Health Management Information System |  |  |

## Discussion

This study found a range of variables from health facility type, managing sector, health management information system characteristics, and location to funding sources that were associated with institutional measurement of population coverage for key health indicators. In general, less than half of primary health care facilities measured population coverage for key health indicators, similar to other low and middle-countries (LMIC) where there is an underperformance of this crucial activity^13^ ascribed to major individual and organizational barriers at frontline health facilities^14^ .

Primary health care institutions that measured population coverage for key health indicators were predominantly community health centers and health centers without bed, and managed mostly by the government or a mixed entity, and located in rural areas. Comparatively, institutions that did not measure population coverage for key health indicators tend to be affiliated with the private for-profit and private non-profit sectors. These data concur with the findings in the literature on LMICs that have been expanding their health system infrastructure to increase access to quality primary health care services delivery in underserved areas^15^. A recent study evidenced the increased proximity (5km) of primary care facilities to residential areas for 91% of the Haitian population, while estimating that slightly more than half of urban residents and merely 8 percent of those residing in rural areas had access to primary care service delivery of good quality^16^. As discovered in other health care systems in LMICs, these major geographical expansions were not always intentionally planned and executed to achieve short and long-term improvements in the quality of health care services delivery and coverage and of health management information systems^15^ in public and private primary health care facilities^1^. In agreement with the literature, most primary health care institutions that measured population coverage for key health indicators had an established health services data collection system, with a reporting frequency of once or more per month, but lacked a corresponding workforce size to manage the health services data and had been experiencing widespread unavailability of health services data technologies. The reverse was paradoxically true for health service data technology availability among primary health care institutions that did not measure population coverage for key health indicators, presumably on account of a plausible primary or exclusive use of such technology for administrative purposes, lack of appropriate HMIS software, and or inadequate personnel.

Although health centers without bed and community health centers comprised a great proportion of primary health care institutions in Haïti, the average outpatient visit for a month was highest at community hospitals. The low proportion of community hospitals to health centers and community health centers, matching the intended role of community hospitals as reference primary health care facilities to ensure continuum of care^10^, might partly explain this high outpatient rate.

The mean outpatient visit was highest at the health facilities that were under mixed management authorities or privately-owned nonprofit. While rural areas had the greatest proportion of primary health care health facilities, the mean outpatient visits for a month was two times higher for health facilities located in urban areas than in rural areas. Similar distribution in outpatient visits has been observed in LMICs and has been attributed to patients’ care seeking-behavior motivated by positive perceptions of quality of care, levels of professionalism, and health facility environments in the private sector^13^, although gaps in these measurements were recently discovered in Haiti and other LMICs^17^. A recent study quantified the quality of health facility infrastructure using a service readiness index in Haiti. It revealed that health facilities scored highest (0.62) in infrastructure quality in urban areas^16^. No significant association was found between infrastructure quality and the volume of patients who utilized prenatal care services. In rural areas, health facility infrastructure quality was positively associated with the volume of patients who utilized antenatal, postnatal and several other care services. In contrast, health facility infrastructure quality exemplified by HMIS measurement performance may not be positively associated with patient volume in rural areas, considering the higher ratio of urban to rural health facilities among institutions that did not measure population coverage for key health indicators compared to those that did. Building or improving the capacity of privately owned urban PHCs in health management information system would necessitate the provision of Incentives and formation of strategic partnerships with national and international stakeholders^1^.

Irrespective of institutional capacity to measure population coverage for key health indicators, a discrepancy was noted between the high prevalence of health services data collection system, data reporting and data completion rate, and the low availability of functional health services data technology. A plausible explanation is the prevailing availability and use of paper-based data collection systems in disparately digitized primary health care facilities^18^ in rural areas^19^ in LMICs. There was an evident shortage of health services data management personnel that was worse in primary health care facilities that did not measure population coverage for key health indicators, which could impact data collection, management, and reporting quality. The Haitian Ministry of Public Health and Population has been implementing the SISNU health management information system built on the DHIS2 platform and providing trainings to stakeholders to solve the complex challenges of data quality collection, management, and reporting^11^. Lessons learned from other health management information system interventions indicate that subpar data quality collection, management and reporting still constitute major barriers to reliable data access and availability in health information systems in LMICs ^20^, which can be remediated through the implementation of interventions of technical, organizational, and behavioral natures. Technical interventions include “improving paper-based data collection forms, using mobile-Health (mHealth) solutions, eHMIS, equipment purchase and maintenance, conducting DQA, improving data storage, and database harmonization”^21^, which have been implemented by the MSPP but not yet adopted by all health facilities^11^. Interventions at the organizational and behavioral levels encompass “training, enhanced training, task-shifting and creation of new roles, supervision, enhanced supervision, engagement of core partners in the intervention, dissemination meetings, incentives, and standardized protocols”^21^, that are reflected to a certain extent in the ongoing efforts by the Ministry of Public health and Population and USAID to build the capacity of the ministry to gain full, impactful, and sustainable governance of the SISNU health information management system^11^. Based on this systematic review of evidence-based interventions, data quality can improve by 80% with these interventions. While electronic health management information system (eHMIS) built on the DHIS2 platform only improves timeliness of data availability and access, the systematic checking and supervision of data quality across the spectrum of data management and health system are sine qua non for ensuring data accuracy and completeness.

Health reforms in LMICs have led to a diversification of funding sources with the concomitant increase in government expenses to impact health outcomes^22^. This study also found a wide variety of funding sources in the health sector in Haiti resulting from the national health care policy on primary health care expansion, access, delivery, and coverage. Out-of-pocket or Individual direct payments constituted the predominant source of funding for institutions that measured population coverage for key health indicators, followed by the Ministry of Health, donor agencies / NGOs, faith-based programs, and international organizations. In contrast, out-of-pockets expenditures significantly decreased with health care system decentralization in most other countries^15^.

Health management information system data collection, management and reporting reliability comes at a cost that can be prohibitive in LMICs. In this study, health facilities are funded primarily by the individual direct payments, ministry of health, donor agencies/NGOs, and faith-based programs. A recent framework has been proposed to strengthen health systems in LMICs. This framework was built under the premises that health financing is contextual based on countries where it is not exclusively under the purview of the technical but also political domains^22^. According to current estimates, scaling up health systems improvements can impact population health outcomes and ultimately lead to the achievement of SDG3 in LMICs, provided that “total health-care spending would increase to a population-weighted mean of $271 per person (range 74–984) across country contexts, and the share of gross domestic product spent on health would increase to a mean of 7.5% (2.1–20.5)”^22^. Based on the authors’ recommendations, the Haitian government ought to incrementally increase their health care spending budget^5,6^ by 1-2%^22^. As a low-income country with a high fragility index score, political stabilization takes precedence and is a cornerstone of health system strengthening^22^.

The results of this study largely reflect the health care system in Haïti. Nevertheless, several limitations were identified. A more recent quinquennial SPA dataset would probably have provided new evidence on the current landscape of the health management information systems and financing structure in primary health care facilities across the 10 departments in Haïti. A time series analysis including the 2013, 2017-2018 and new SPA data would have permitted a longitudinal analysis of profile change in the health system characteristics, infrastructure, and financing in Haïti. Additional technical, behavioral, and organizational determinants of health facility performance^21,23^ could have been statistically significantly associated with the capacity of institutions to perform population coverage measurement for key health indicators but were not included in this study. Expert verification of health services data completeness was not performed to avoid overestimation. The SPA survey respondents may have misconstrued the definition of population coverage for key health indicators to either reflect the patient population or population in the catchment area of the primary health care facilities, which potentially introduced bias in this study data analysis. However, it is likely that this bias is non-differential, and therefore the observed odds ratios may be conservatively lower than the actual estimates.

## Conclusion

This study highlighted the profile and associations of health facilities characteristics, infrastructure, and funding sources according to their performance level in measurement of population coverage for key health indicators in Haïti. Institutions that measured population health coverage were predominantly community health centers located in rural areas and managed by the government, and in more than half of cases unequipped with a functional health management information system and lacking adequate health service data management personnel. These institutions were predominantly funded by the Ministry of Health, out-of-pocket payments, donor agencies/NGOs, faith-based programs, and international organizations. These data and the findings on the SISNU or HMIS platform implementation outcomes substantiate the continued need for evidence-based and contextual technical, organizational, behavioral, and financial interventions at the local and national levels to optimize measurement activities of population coverage for key health indicators in Haïti.

## Data Availability

The data can be made available upon request or accessed through the DHS platform.

## Appendix

**Table A.** Mean Outpatient Visits in The Previous Month by Health Facility characteristics in Haïti.

|  |  |  |  |
| --- | --- | --- | --- |
| General Characteristics | Mean Outpatient Visits in The Previous Month |  |  |
|  | Mean | Standard Error | [95% Confidence Interval] |
| **Health Facility Type** |  |  |  |
| Community Hospitals | 1045 | 128 | [795, 1296] |
| Health Centers with Bed | 412 | 46 | [322, 503] |
| Health Centers without Bed | 445 | 48 | [351, 538] |
| Community Health Centers | 175 | 17 | [143, 208] |
| **Health Facility Sector** |  |  |  |
| Government | 376 | 43 | [293, 460] |
| Private Non-Profit | 467 | 78 | [314, 620] |
| Private For Profit | 392 | 44 | [305, 478] |
| Mixed | 470 | 59 | [355, 585] |
| **Health Facility Location** |  |  |  |
| Urban | 562 | 54 | [457, 668] |
| Rural | 329 | 27 | [277, 381] |
| **Health Facility Activity** |  |  |  |
| Measures Population Coverage for Key Indicators | 312 | 30 | [254, 369] |
| No Measure of Population Coverage for Key Indicators | 494 | 40 | [415, 573] |
| Weighted n and % P Value: Rao and Scott second-order corrected Chi-square |  |  |  |

**Table B.**
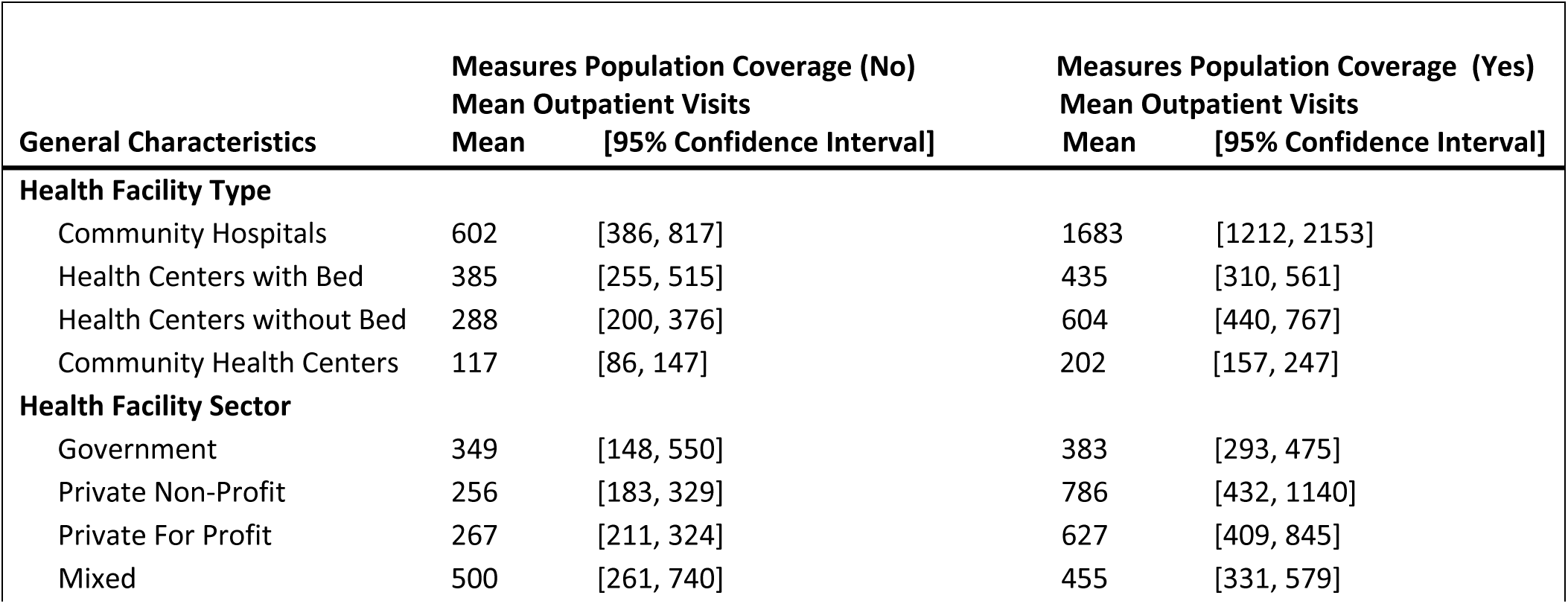
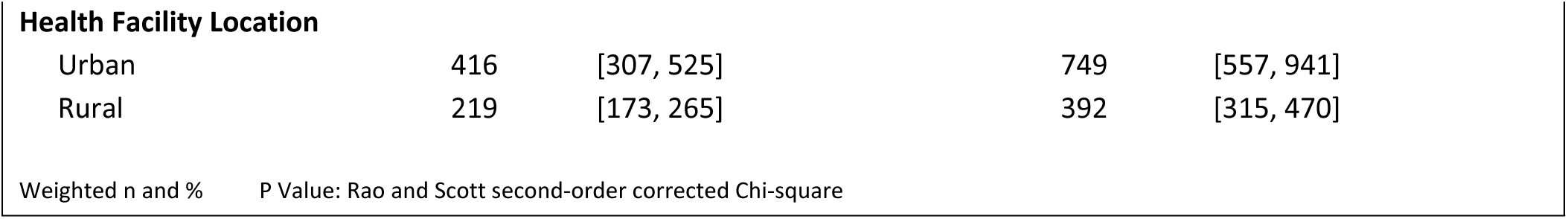
Mean Outpatient Visits in The Previous Month Per Health Facility Characteristic according to Institutional Measurement Status of Population Coverage for Key Health Indicators in Haïti.

|  |  |  |  |  |
| --- | --- | --- | --- | --- |
| General Characteristics | Measures Population Coverage (No)<br>Mean Outpatient Visits |  | Measures Population Coverage (Yes)<br>Mean Outpatient Visits |  |
|  | Mean | [95% Confidence Interval] | Mean | [95% Confidence Interval] |
| **Health Facility Type** |  |  |  |  |
| Community Hospitals | 602 | [386, 817] | 1683 | [1212, 2153] |
| Health Centers with Bed | 385 | [255, 515] | 435 | [310, 561] |
| Health Centers without Bed | 288 | [200, 376] | 604 | [440, 767] |
| Community Health Centers | 117 | [86, 147] | 202 | [157, 247] |
| **Health Facility Sector** |  |  |  |  |
| Government | 349 | [148, 550] | 383 | [293, 475] |
| Private Non-Profit | 256 | [183, 329] | 786 | [432, 1140] |
| Private For Profit | 267 | [211, 324] | 627 | [409, 845] |
| Mixed | 500 | [261, 740] | 455 | [331, 579] |
| **Health Facility Location** |  |  |  |  |
| Urban | 416 | [307, 525] | 749 | [557, 941] |
| Rural | 219 | [173, 265] | 392 | [315, 470] |

Weighted n and % P Value: Rao and Scott second-order corrected Chi-square

**Table C.**
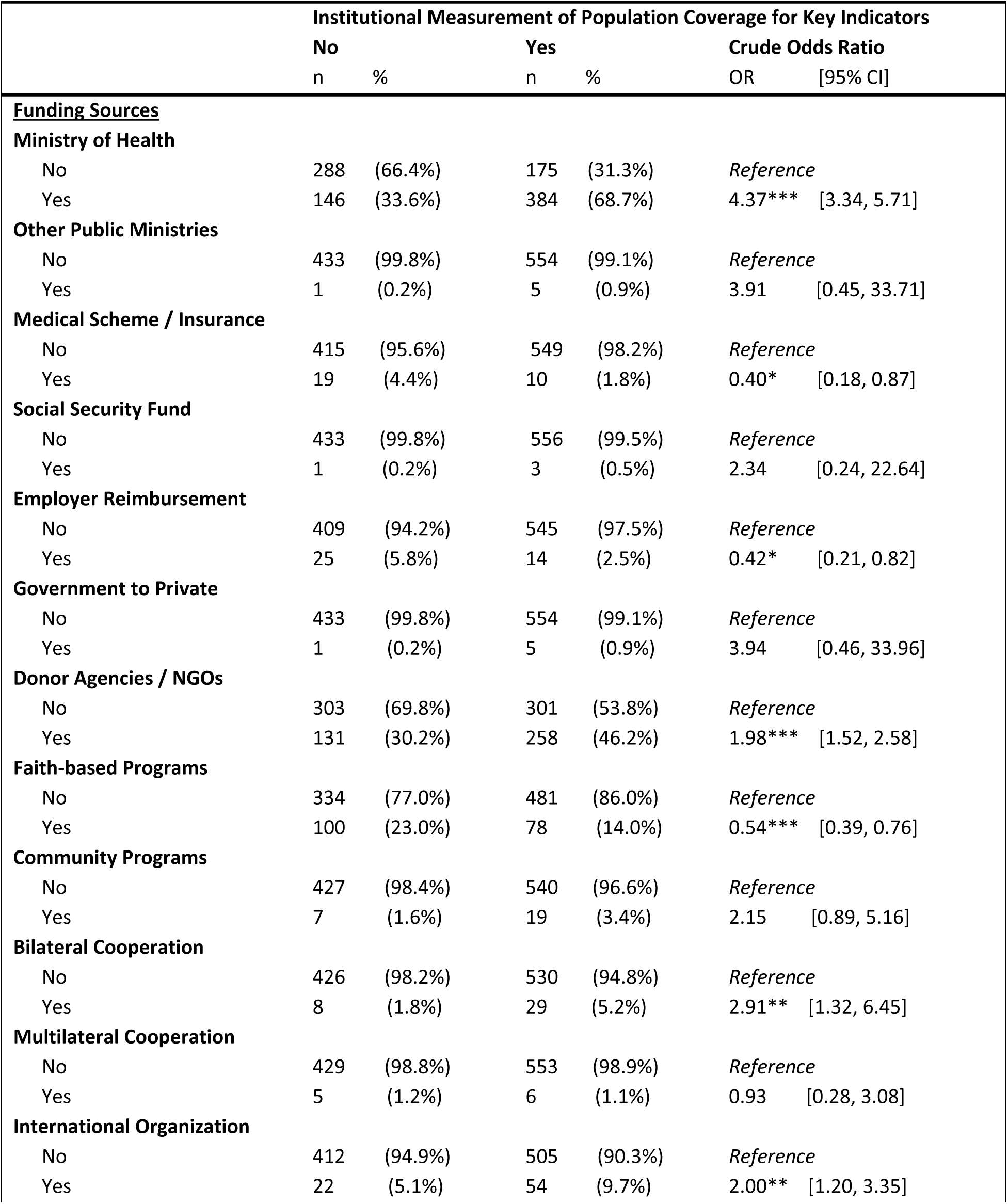
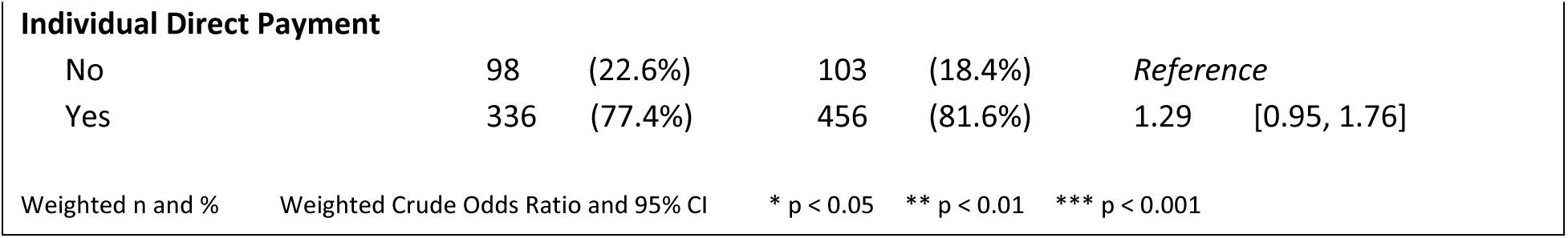
Association of Primary Health Care Facility Funding Sources with Institutional Measurement of Population Coverage for Key Health Indicators in Haïti.

## References

1. Kruk ME, Gage AD, Arsenault C, et al. High-quality health systems in the sustainable development goals era: Time for a revolution. The Lancet global health. 2018;6(11):e1196–e1252. doi: 10.1016/S2214-109X(18)30386-3.

2. Moreno-Serra R, Anaya-Montes M, Smith PC. Potential determinants of health system efficiency: Evidence from latin america and the caribbean. PloS one. 2019;14(5):e0216620. doi: 10.1371/journal.pone.0216620.

3. Akseer N, Phillips DE. Drivers of success in global health outcomes: A content analysis of exemplar studies. PLOS global public health. 2024;4(5):e0003000. doi: 10.1371/journal.pgph.0003000.

4. Abdi A, Abdulkader RS, Adetokunboh OO, et al. Measuring progress from 1990 to 2017 and projecting attainment to 2030 of the health-related sustainable development goals for 195 countries and territories: A systematic analysis for the global burden of disease study 2017. The Lancet. 2018;392(10159):2091–2138. doi: 10.1016/S0140-6736(18)32281-5.

5. World Health Organization. Data indicators: Haïti. . https://data.who.int/countries/332. Updated 2023.

6. World Bank Group: Data. Current health expenditure per capita (current US$). https://data.worldbank.org/indicator/SH.XPD.CHEX.PC.CD. Updated 2023. Accessed July 17, 2024.

7. Institut Haïtien de l’Enfance (IHE) et ICF International. Évaluation de la prestation des services de soins de santé 2017-2018. https://dhsprogram.com/pubs/pdf/SPA29/SPA29.pdf. Accessed July 17, 2024.

8. Sanhueza A, Carvajal-Vélez L, Mújica OJ, Vidaletti LP, Victora CG, Barros AJ. SDG3-related inequalities in women’s, children’s and adolescents’ health: An SDG monitoring baseline for latin america and the caribbean using national cross-sectional surveys. BMJ open. 2021;11(8):e047779. doi: 10.1136/bmjopen-2020-047779.

9. Fene F, Ríos-Blancas MJ, Lachaud J, et al. Life expectancy, death, and disability in haiti, 1990-2017: A systematic analysis from the global burden of disease study 2017. Revista panamericana de salud pública. 2020;44(136):e136. doi: 10.26633/RPSP.2020.136.

10. Ministère de la Santé Publique et de la Population. Représentation du système de santé. . 2015. https://mspp.gouv.ht/site/downloads/Paquet_minimum_de_services_1er%20niveau.pdf.

11. USAID. Haïti strategic health information system (HIS) project. 2021. https://pdf.usaid.gov/pdf_docs/PA00Z4GM.pdf.

12. StataCorp. Stata statistical software: Release 18. https://www.stata.com/. Updated 2023. Accessed July 17, 2024.

13. Basu S, Andrews J, Kishore S, Panjabi R, Stuckler D. Comparative performance of private and public healthcare systems in low- and middle-income countries: A systematic review. PLoS medicine. 2012;9(6):e1001244. doi: 10.1371/journal.pmed.1001244.

14. Li, M., Brodsky, I., & Geers, E.&nbsp, . Barriers to use of health data in low-and middle-income countries—A review of the literature. measure evaluation. *Carolina Population Center*, University of North Carolina at Chapel Hill. 2018:1–29.

15. Cobos Muñoz D, Merino Amador P, Monzon Llamas L, Martinez Hernandez D, Santos Sancho JM. Decentralization of health systems in low and middle income countries: A systematic review. Int J Public Health. 2017;62(2):219–229. doi: 10.1007/s00038-016-0872-2.

16. Gage AD, Leslie HH, Bitton A, et al. Does quality influence utilization of primary health care? evidence from haiti. Globalization and health. 2018;14(1):59. doi: 10.1186/s12992-018-0379-0.

17. Macarayan EK, Gage AD, Doubova SV, et al. Assessment of quality of primary care with facility surveys: A descriptive analysis in ten low-income and middle-income countries. The Lancet global health. 2018;6(11):e1176–e1185. doi: 10.1016/S2214-109X(18)30440-6.

18. Kaboré SS, Ngangue P, Soubeiga D, et al. Barriers and facilitators for the sustainability of digital health interventions in low and middle-income countries: A systematic review. Frontiers in digital health. 2022;4:1014375. doi: 10.3389/fdgth.2022.1014375.

19. Maita, K. C., Maniaci, M. J., Haider, C. R., Avila, F. R., Torres-Guzman, R. A., Borna, S., Lunde, J. J., Coffey, J. D., Demaerschalk, B. M., & Forte, A. J. The impact of digital health solutions on bridging the health care gap in rural areas: A scoping review. The Permanente journal. 2024. doi: 10.7812/TPP/23.134.

20. Rendell N, Lokuge K, Rosewell A, Field E. Factors that influence data use to improve health service delivery in low- and middle-income countries. Global health science and practice. 2020;8(3):566–581. doi: 10.9745/GHSP-D-19-00388.

21. Lee J, Lynch CA, Hashiguchi LO, et al. Interventions to improve district-level routine health data in low-income and middle-income countries: A systematic review. BMJ global health. 2021;6(6):e004223. doi: 10.1136/bmjgh-2020-004223.

22. Stenberg K, Hanssen O, Edejer TT, et al. Financing transformative health systems towards achievement of the health sustainable development goals: A model for projected resource needs in 67 low-income and middle-income countries. The Lancet global health. 2017;5(9):e875–e887. doi: 10.1016/S2214-109X(17)30263-2.

23. Klesta Hoxha, Yuen W Hung, Bridget R Irwin, Karen A Grepin. Understanding the challenges associated with the use of data from routine health information systems in low- and middle-income countries: A systematic review. Health information management : journal of the Health Information Management Association of Australia. 2022;51(3):135–148. http://search.informit.org/doi/10.3316/informit.641751128511242.

